# Feasibility of Estimating Cardiac Indices Using Cardiac Surgery Anesthesia Records in a Multicenter Cohort

**DOI:** 10.1101/2025.08.28.25333855

**Authors:** Emily A Balczewski, Graciela Mentz, MPOG Collaborators, Karandeep Singh, Michael Mathis

**Affiliations:** Medical Scientist Training Program, University of Michigan Medical School, Ann Arbor, MI, USA; Department of Computational Medicine and Bioinformatics, University of Michigan Medical School, Ann Arbor, MI, USA; Department of Anesthesiology, University of Michigan Medical School, Ann Arbor, MI, USA; Joan and Irwin Jacobs Center for Health Innovation, University of California, San Diego, CA, USA; Department of Medicine - Division of Biomedical Informatics, University of California, San Diego, CA, USA

## Abstract

**Purpose:** Cardiac index (CI) is a key physiologic indicator correlated with end-organ perfusion in cardiac surgical patients, yet it is not routinely measured in all cases. This study evaluated the accuracy of estimating CI using routinely available physiologic monitor data, adjusted for relevant patient, physiologic, and procedural factors documented in perioperative anesthesia records.

**Methods:** We analyzed anesthesia records from adult cardiac surgical patients with thermodilution-based CI measurements across seven US institutions from 2014 to 2022. Four published formulas—based on intraoperative blood pressure and heart rate—were used to estimate CI in generalized linear models, with adjustment for perioperative patient and procedure characteristics. Bland-Altman analysis compared adjusted CI estimates to reference thermodilution CI values. The ability of each estimator to classify patients with low CI (<2.2 L/min/m^2^) was assessed for concordance.

**Results:** In a cohort of 5,989 patients, the median (IQR = interquartile range) thermodilution-based CIs were 2.1 (1.8–2.6) and 2.4 (2.0–2.9) L/min/m^2^ before and after cardiopulmonary bypass, respectively. The best-performing formula, Liljestrand and Zander, achieved mean absolute errors of 0.45 and 0.47 L/min/m^2^ before and after bypass, respectively. However, its reliability in classifying low CI was limited (Cohen’s kappa = 0.26 pre-bypass, 0.20 post-bypass).

**Conclusion:** Routinely collected physiologic and patient data can be used to generate population-level cardiac index estimates in adult cardiac surgery patients when appropriately adjusted, though individual-level discrimination of low CI is limited. These findings inform future large-scale perioperative hemodynamic research.

## Introduction

Cardiac indices (CIs) are important, modifiable physiologic measures correlated to end-organ perfusion – and complications resulting from organ malperfusion – among patients undergoing cardiac surgery [1]. Nonetheless, benefits of directly measuring CI through invasive indicator dilution techniques, such as thermodilution via a pulmonary artery catheter (PAC), are counterbalanced by risks associated with placement of such monitors (e.g., arrhythmia, thrombus formation, infection, pulmonary artery rupture) [2]. As a result, approximately only one-third of adult cardiac surgical patients in the US receive PAC monitoring [3], and multiple methods to ascertain CI via less invasive measures have been developed [4], [5].

While some of these techniques show clinical promise, most rely on proprietary physiological sensors or arterial waveforms not routinely captured in the anesthesia record, with variable uptake across institutions [6]. Conversely, studies which instead use routinely-collected physiologic data to estimate CI have largely shown limited reliability [7], [8], [9], [10], [11], [12], [13], [14]; one study in a small cohort of patients undergoing coronary artery bypass graft (CABG) surgery reported a 49% error rate for its highest-performing estimator (Liljestrand and Zander) [15]. The limited performance of these simple physiologic estimators is potentially explained by the complexity of physiology during cardiac surgery, with measurement variation attributable to patient demographics, concurrent cardiac conditions, cardiovascular medication administration, and surgical interventions [16], [17], [18]. Among studies of CI estimators, few have attempted to control for such factors, although one found that inclusion of age and sex significantly improved the performance of cardiac output (CO) estimates in a single center [7].

Nonetheless, whereas physiologic estimators of CI have largely unproven utility to inform clinical care decisions of individual patients, the usefulness of such estimators for population-level observational research remains unresolved. Specifically, opportunities may exist to explore clinical care questions leveraging existing electronic health record (EHR) and surgical registry data to understand fundamental relationships between low CI and malperfusion-related outcomes. In such study designs, CI estimators – if found to have at least moderate accuracy at the population level – may aid in uncovering relationships between CI-modifying hemodynamic management interventions and cardiac surgical outcomes mediated by low CI, using routinely collected clinical data among the majority of patients for whom direct CI measurements are not obtained.

To address the limitations of previous studies and explore this line of clinical scientific inquiry, we performed this multicenter observational study of adult cardiac surgical patients with invasive CI measurements undergoing a variety of procedures. We hypothesized that simple physiologic CI estimators were associated with invasive, thermodilution-based CI measurements when adjusted for relevant perioperative patient and procedure information routinely available within electronic anesthesia records and surgical registries.

## Methods

### Study Design

This study was deemed exempt by the Michigan IRBMED (HUM00206345). The inclusion/exclusion criteria, data collection/cleaning methods, and statistical approaches of the study were approved a priori within a peer-review forum [19]. The study manuscript adheres to the REporting of studies Conducted using Observational Routinely-collected health Data (RECORD) extension of the Strengthening the Reporting of Observational Studies in Epidemiology (STROBE) guidelines [20].

### Data Source

Data were obtained from the Multicenter Perioperative Outcomes Group (MPOG) and Society of Thoracic Surgeons Adult Cardiac Surgical Database (STS-ACSD) registries. Procedures for acquiring, validating, and transferring data from participating sites to MPOG and STS-ACSD have been previously described [21], [22], [23]. Additionally, MPOG and STS-ACSD handled missing or invalid data according to registry-specific protocols [24], [25]. Data from seven US hospitals participating in both MPOG and STS-ACSD were integrated locally for this study.

### Study Population

Adult patients undergoing cardiac surgery from January 1, 2014 to February 1, 2022 with at least one intraoperative CI measurement were included. Cardiac surgical procedures included coronary artery bypass grafts (CABG), valve repairs or replacements, thoracic aortic procedures, or combinations thereof. Among patients who underwent multiple procedures meeting inclusion criteria during the study period, the index case was used. A complete case analysis was performed, due to low rates of missingness among model covariates.

### Primary Outcome - Thermodilution-based Cardiac Index

The primary outcome was a CI reference standard, measured via a PAC thermodilution technique. Measurements were reported within the intraoperative anesthesia record at each institution either as a CI or cardiac output (CO) value. CO measurements obtained from these sources were converted to CI measurements by normalizing according to calculated body-surface area [26], [27]. Each case was randomly downsampled to two intraoperative CI measurements: one before cardiopulmonary bypass [pre-CPB] and one after [post-CPB] when available, or a single CI measurement when only pre-CPB or post-CPB measurements were available. CI measurements were assessed for normality, symmetry, and potential outliers with histograms, Q-Q plots, violin plots, and descriptive statistics.

### Exposure Variable - CI Estimators

Four previously reported estimators of CI were computed from routinely available intraoperative physiologic data: systolic blood pressure (SBP), mean arterial pressure (MAP), diastolic blood pressure (DBP), and heart rate (HR), normalized by body surface area as analogous to CO versus CI measurements (Table 1). Measurements of these hemodynamic factors were obtained during the same minute of cardiac index measurement. Where multiple measurements were found in a single minute, the median value was used. Rarely, when no hemodynamic measurements were obtained that minute, the most recent measurement was carried forward from a previous minute.

**Table 1.** Evaluated Estimators of Cardiac Index. CI = cardiac index; SBP = systolic blood pressure; DBP = diastolic blood pressure; MAP = mean arterial pressure; HR = heart rate; BSA = body surface area.

| Estimator of CI | Formula |
| --- | --- |
| Mean Arterial Pressure (MAP) | $(1/3 \text{ SBP} + 2/3 \text{ DBP}) / \text{BSA}$ |
| Windkessel [28] | $(\text{SBP} - \text{DBP}) * \text{HR} / \text{BSA}$ |
| Herd [29] | $(\text{MAP} - \text{DBP}) * \text{HR} / \text{BSA}$ |
| Liljestrand and Zander [10] | $(\text{SBP} - \text{DBP}) / (\text{SBP} + \text{DBP}) * \text{HR} / \text{BSA}$ |

### Adjustment Variables - Patient, Case, and Intraoperative Factors

To adjust for variables potentially influencing the relationship between CI estimators and the invasive thermodilution-based CI reference standard, additional control variables included (i) patient baseline characteristics (age, sex, race, preoperative left ventricular ejection fraction, the presence of cardiogenic shock before surgery, and type of surgical procedure(s)), (ii) intraoperative hemodynamic factors (central venous pressure, O2 saturation), and (iii) intraoperative vasoactive-inotropic score (VIS) [30]. Intraoperative hemodynamic factors and current doses of vasopressors and inotropes (for VIS) were obtained in the same minute of CI measurement and processed analogously to CI estimators. Preoperative patient and surgical procedure data were collected via the STS-ACSD; additional pre- and intraoperative data were acquired from MPOG using validated, open-source phenotypes [24]. Additionally, to account for possible clinician- and hospital-level variation in PAC measurement and documentation practices, multivariable adjustment was performed using a multilevel modelling structure, with surgical procedures nested within anesthesiologists nested within institutions.

### Statistical Analyses

Statistical analyses were performed using SAS version 9.4 (SAS Institute, USA) and R version 4.2.1 (R Foundation for Statistical Computing, Vienna, Austria). Model selection followed a multi-criteria approach, using a combination of clinical expertise and empirical selection with least absolute shrinkage and selection operator (LASSO). A multilevel analysis approach was selected after evaluating the modified intraclass correlation estimates of hospital and primary anesthesia attending on the total variation of CI. Both levels were retained for analysis after >1% of the total variance were explained within each level. Generalized linear mixed models were constructed to compute an adjusted estimated CI using the estimators in Table 1, adjusted for pre- and intraoperative patient and case factors. The final model specification was as follows:

Adjusted CI = CI estimator + (patient- and case-level covariates) + (1 | hospital) + (1 | primary anesthesia attending) + error

A Bland-Altman analysis was performed for each model-adjusted CI estimator using the 95% confidence interval of the limits of agreement (±2.093√ 3/n^*^s; s = is the standard deviation of the bias; n = sample size) [31]. Performance of the model-produced adjusted CI estimates were additionally assessed with an error analysis that included mean absolute percentage error

(MAPE) and mean absolute error (MAE) [32]. Finally, the ability of the adjusted CI estimates to discriminate between clinically low CI (<2.2 L/min/m^2^) versus normal or high CI was assessed with sensitivity, specificity, positive predictive value (PPV), negative predictive value (NPV), and Cohen’s kappa [33].

## Results

Of the 24,726 cardiac surgical cases eligible for inclusion, 7,169 (30.0%) contained at least one invasive thermodilution-based CI measurement, among which 5,989 (83.7%) contained complete multivariable data for estimating CI values across 7 US hospitals (Table 2; Supplemental Table 1 in <u>Online Resource 1</u>). The cohort included 30.4% female and 85.3% White patients with a median age of 65 years (interquartile range (IQR) = [55-72]). Non-mutually exclusive procedures included 68.7% valve repair/replacement, 32.6% CABG, and 26.0% thoracic aortic procedures. Additionally, the analysis cohort included patients with variable preoperative cardiac function (LVEF median 58%, IQR = [45-65]), including 4.0% of patients with surgical registry-defined preoperative cardiogenic shock.

**Table 2.**
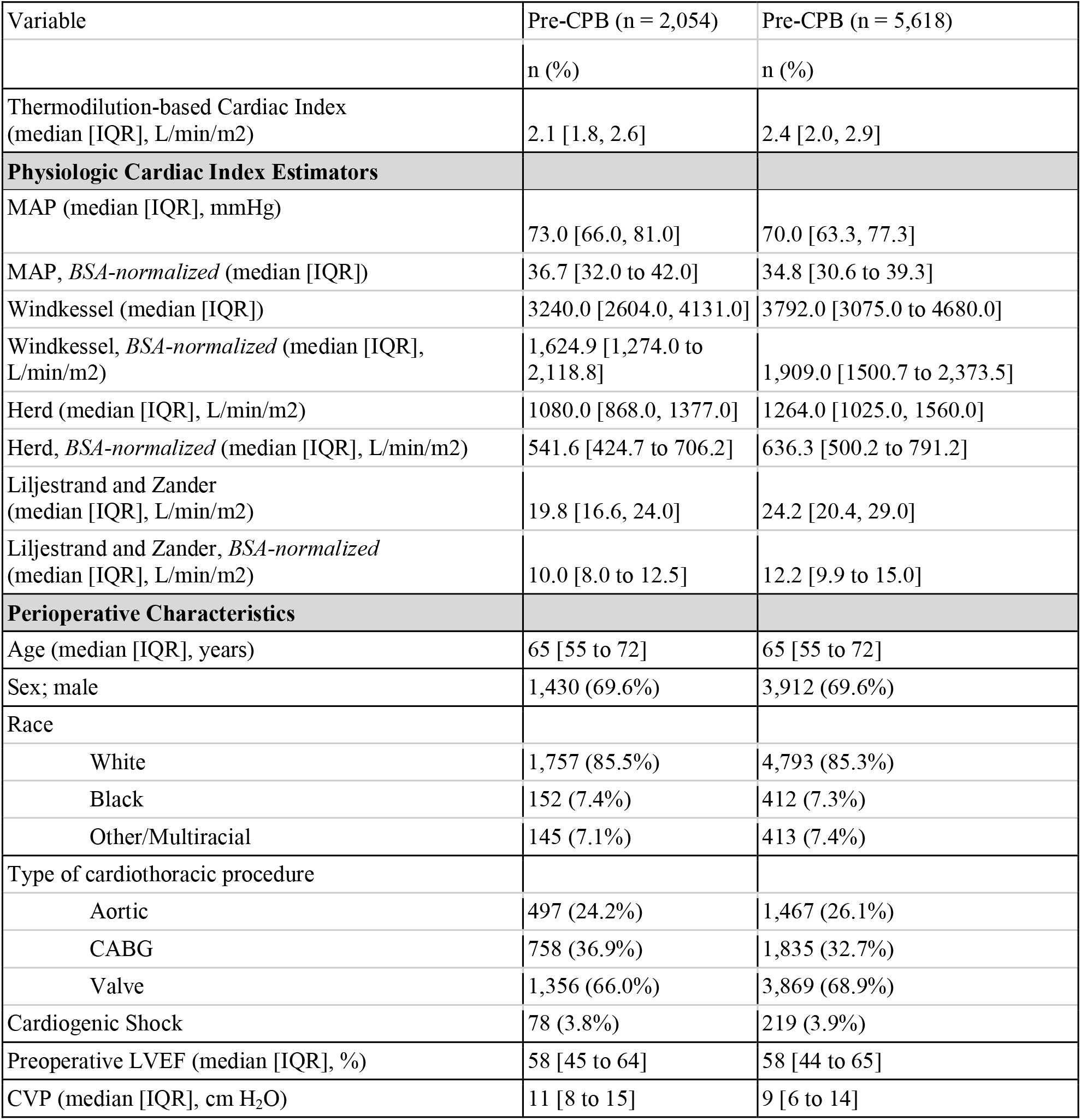
Pre- and post-CPB cohort characteristics. CPB = cardiopulmonary bypass; IQR = interquartile range; BSA = body surface area; MAP = mean arterial pressure; CABG = coronary artery bypass graft; LVEF = left ventricular ejection fraction; CVP = central venous pressure.

| Variable | Pre-CPB (n = 2,054) | Pre-CPB (n = 5,618) |
| --- | --- | --- |
|  | n (%) | n (%) |
| Thermodilution-based Cardiac Index (median [IQR], L/min/m<sup>2</sup>) | 2.1 [1.8, 2.6] | 2.4 [2.0, 2.9] |
| **Physiologic Cardiac Index Estimators** |  |  |
| MAP (median [IQR], mmHg) | 73.0 [66.0, 81.0] | 70.0 [63.3, 77.3] |
| MAP, *BSA-normalized* (median [IQR]) | 36.7 [32.0 to 42.0] | 34.8 [30.6 to 39.3] |
| Windkessel (median [IQR]) | 3240.0 [2604.0, 4131.0] | 3792.0 [3075.0 to 4680.0] |
| Windkessel, *BSA-normalized* (median [IQR], L/min/m<sup>2</sup>) | 1,624.9 [1,274.0 to 2,118.8] | 1,909.0 [1500.7 to 2,373.5] |
| Herd (median [IQR], L/min/m<sup>2</sup>) | 1080.0 [868.0, 1377.0] | 1264.0 [1025.0, 1560.0] |
| Herd, *BSA-normalized* (median [IQR], L/min/m<sup>2</sup>) | 541.6 [424.7 to 706.2] | 636.3 [500.2 to 791.2] |
| Liljestrand and Zander (median [IQR], L/min/m<sup>2</sup>) | 19.8 [16.6, 24.0] | 24.2 [20.4, 29.0] |
| Liljestrand and Zander, *BSA-normalized* (median [IQR], L/min/m<sup>2</sup>) | 10.0 [8.0 to 12.5] | 12.2 [9.9 to 15.0] |
| **Perioperative Characteristics** |  |  |
| Age (median [IQR], years) | 65 [55 to 72] | 65 [55 to 72] |
| Sex; male | 1,430 (69.6%) | 3,912 (69.6%) |
| Race |  |  |
| White | 1,757 (85.5%) | 4,793 (85.3%) |
| Black | 152 (7.4%) | 412 (7.3%) |
| Other/Multiracial | 145 (7.1%) | 413 (7.4%) |
| Type of cardiothoracic procedure |  |  |
| Aortic | 497 (24.2%) | 1,467 (26.1%) |
| CABG | 758 (36.9%) | 1,835 (32.7%) |
| Valve | 1,356 (66.0%) | 3,869 (68.9%) |
| Cardiogenic Shock | 78 (3.8%) | 219 (3.9%) |
| Preoperative LVEF (median [IQR], %) | 58 [45 to 64] | 58 [44 to 65] |
| CVP (median [IQR], cm H<sub>2</sub>O) | 11 [8 to 15] | 9 [6 to 14] |

Among cases with complete data, 2,054 (34.3%) included CI values prior to CPB and 5,618 (93.8%; non-mutually exclusive) after CPB (Table 2; Supplemental Table 1 in <u>Online Resource 1</u>). Following adjustment for perioperative factors, CI models converged for all four estimators and improved upon the null model; full model results can be found in <u>Online Resource 2</u>.

Notably, the Windkessel and Herd estimators were perfectly correlated (Spearman correlation = 1.00) with each other and highly correlated (0.89) with the Liljestrand and Zander estimator and, therefore, produced similar model results (full correlation matrix in Supplemental Table 2 in <u>Online Resource 1</u>).

Three of four estimators (excluding MAP) were statistically significant (p < 0.05 by unpaired t-test) in all pre- and post-CPB models (<u>Online Resource 2</u>). Significant covariates across all pre- and post-CPB models included: age, sex, race, preoperative LVEF, and VIS. Significant covariates in only the post-CPB models included aortic procedure type. Significant covariates in only the pre-CPB models included presence of preoperative cardiogenic shock and CABG procedure type (excluding MAP estimator model).

Hospital explained 8.0-9.6% of the total variation in CI measurements across all models and primary anesthesia attending explained up to a further 2.3% of the variation.

Bland-Altman and error analyses are detailed in Table 3. Adjusted estimated CIs were similar across all four models. Models showed a modest MAPE (pre-CPB range 16.2-16.8%; post-CPB range 15.4-15.7%) and MAE (pre-CPB range 0.45-0.47 L/min/m^2^; post-CPB range 0.47-0.49 L/min/m^2^). Spearman correlations with the thermodilution-based CI reference standard were weak to moderate (pre-CPB range 0.31-0.38; post-CPB range 0.38-0.44). Overall, the Liljestrand and Zander estimator had the strongest performance across three of four comparison measures. The Bland-Altman plot for Liljestrand and Zander can be found in Figure 1 and the remaining estimators in Supplemental Figure 1 in <u>Online Resource 1</u>.

**Table 3.** Bland-Altman and error analysis. Best performing values for each comparison metric highlighted in gray. CPB = cardiopulmonary bypass; SD = standard deviation.

| Adjusted Estimated CI Model | MAP Model |  | Windkessel Model |  | Herd Model |  | Liljestrand and Zander Model |  |
| --- | --- | --- | --- | --- | --- | --- | --- | --- |
|  | Pre-CPB | Post-CPB | Pre-CPB | Post-CPB | Pre-CPB | Post-CPB | Pre-CPB | Post-CPB |
| Median [IQR], L/min/m<sup>2</sup> | 2.20<br>[2.1-2.4] | 2.50<br>[2.3-2.7] | 2.21<br>[2.0-2.4] | 2.50<br>[2.3-2.7] | 2.21<br>[2.0-2.4] | 2.50<br>[2.3-2.7] | 2.20<br>[2.0-2.4] | 2.50<br>[2.3-2.7] |
| Mean Absolute Percent Error (%) | 16.8 | 15.7 | 16.5 | 15.6 | 16.5 | 15.5 | 16.2 | 15.4 |
| Mean Absolute Error (L/min/m<sup>2</sup>) | 0.47 | 0.49 | 0.46 | 0.48 | 0.46 | 0.48 | 0.45 | 0.47 |
| Spearman Correlation | 0.31 | 0.38 | 0.36 | 0.42 | 0.31 | 0.40 | 0.38 | 0.44 |

**Figure 1.**
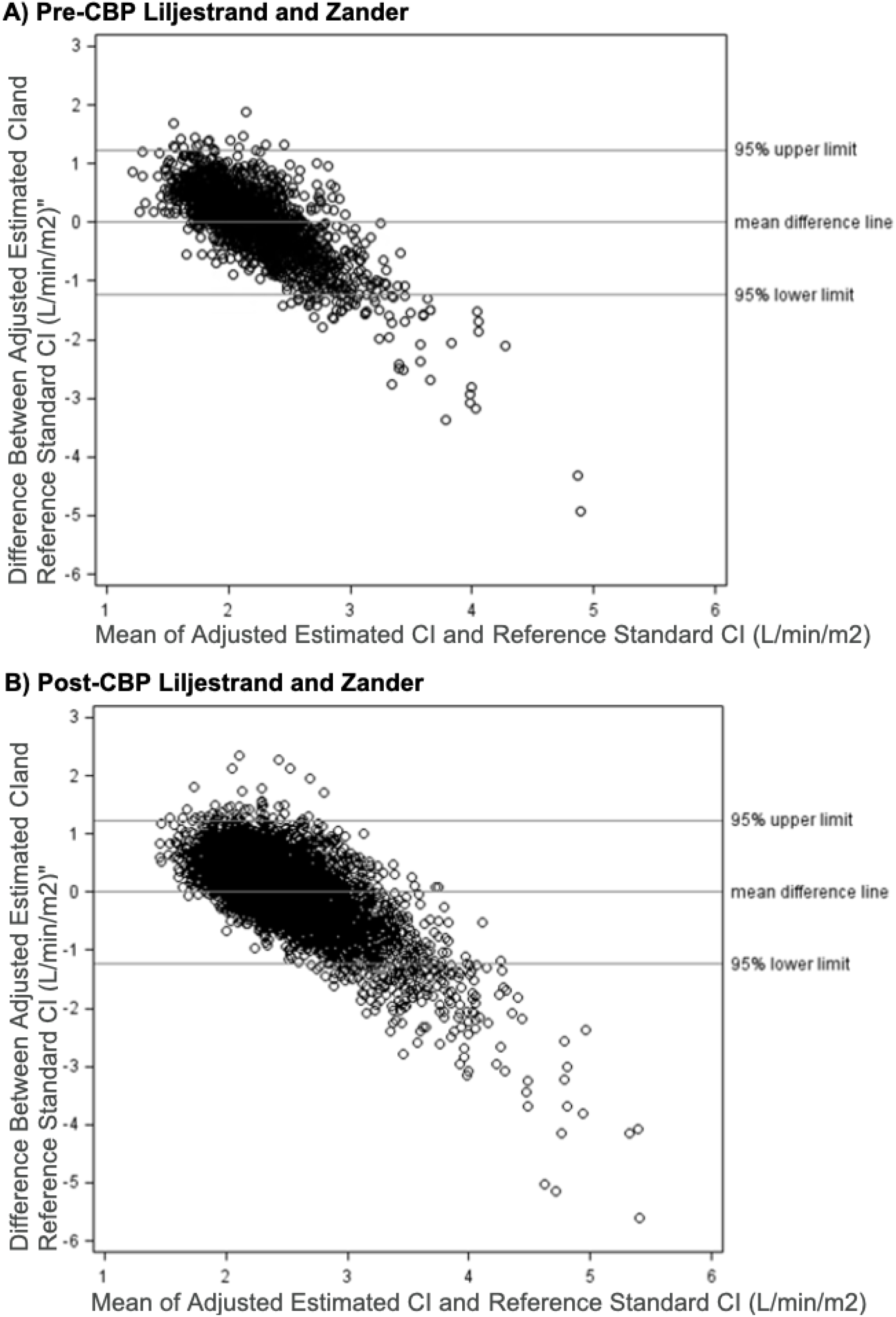
Bland-Altman plots for the Liljestrand and Zander estimator pre- and post-CPB. CPB = cardiopulmonary bypass.

Validity of CI estimates at discriminating patients with low CI (<2.2 L/min/m^2^) was assessed (Table 4). Confusion matrices for each model can be found in Supplemental Table 3 in <u>Online Resource 1</u>. Agreement between CI estimates and CI measurements was low (kappa <0.4) across all models pre- and post-CPB. Similarly to the Bland-Altman and error analysis, the Liljestrand and Zander estimator had marginally better performance than other estimators in classifying patients as having low CI.

**Table 4.** Agreement of true CI and estimated CI for low CI <2.2 L/min/m^2^. Best performing values for each comparison metric highlighted in gray. CPB = cardiopulmonary bypass; PPV = positive predictive value; NPV = negative predictive value.

|  | MAP |  | Windkessel |  | Herd |  | Liljestrand and Zander |  |
| --- | --- | --- | --- | --- | --- | --- | --- | --- |
|  | Pre-CPB | Post-CPB | Pre-CPB | Post-CPB | Pre-CPB | Post-CPB | Pre-CPB | Post-CPB |
| Sensitivity | 0.57 | 0.24 | 0.61 | 0.28 | 0.61 | 0.28 | 0.62 | 0.31 |
| Specificity | 0.66 | 0.89 | 0.65 | 0.88 | 0.65 | 0.88 | 0.65 | 0.87 |
| PPV | 0.67 | 0.55 | 0.68 | 0.58 | 0.68 | 0.58 | 0.68 | 0.59 |
| NPV | 0.56 | 0.67 | 0.58 | 0.68 | 0.58 | 0.68 | 0.58 | 0.68 |
| Cohen's Kappa | 0.23 | 0.14 | 0.26 | 0.185 | 0.26 | 0.19 | 0.26 | 0.20 |

## Discussion

In this multicenter observational study of nearly 6,000 adult cardiac surgical patients across seven US hospitals, we evaluated the feasibility of estimating thermodilution-based CI measurements using data routinely available from electronic anesthesia records, combined with surgical registry data. We report that four different standard physiologic monitor-based estimators are independently associated with the PAC thermodilution-based CI reference standard, among models that adjust for clinically relevant patient and procedure characteristics. Our findings have several important clinical and research implications.

First, although our models achieved modest accuracy in estimating CI, as evidenced by MAE and MAPE values which were lower than commonly cited difference detection thresholds (20– 26%) between serial thermodilution measurements [34], their clinical utility for informing individual patient care decisions is limited. The estimates consistently showed underestimation at higher CI values and overestimation at lower values, as illustrated in the Bland-Altman analyses. Consequently, as supported by the poor agreement measured via Cohen’s kappa, these estimators are not sufficiently reliable for discriminating between patients with low versus normal CI—a finding with important implications among cardiac surgical patients for whom the identification and management of low CI is potentially critical to outcomes [35]. Thus, while our study shows that minimally invasive CI estimation methods drawn from routinely available EHR data are feasible, it also reinforces that their current performance does not support replacement of direct measurement in individual patient management during cardiac surgery.

Second, whereas the physiologic monitor-based estimators may have limited roles in informing clinical decisions at the bedside, such CI estimators derived from routinely available EHR and surgical registry data may remain valuable for research purposes, particularly at the population level. The modest accuracy and similar error rates across CI estimator models suggest that, when adjusted with appropriate patient, procedural, and intraoperative variables, estimators offer a practical solution for estimating CI within large-scale datasets. This is particularly relevant for epidemiologic questions exploring CI as a mediator of adverse outcomes in cardiac surgery, where direct measurements of CI are uncommonly available for the majority of patients. Importantly, our multivariable models benefitted from inclusion of expert-selected covariates and the integration of multicenter and registry data, leading to improved performance over simpler models. Factors such as preoperative cardiogenic shock—a variable often excluded in previous investigations due to the exclusion of unstable patients—were independently associated with CI in our study, highlighting the advantage of our comprehensive and inclusive modeling approach. These findings support the consideration of patient demographic and case-mix variables in future studies seeking to generate or apply CI estimates from real-world EHR data.

Third, when comparing the evaluated physiologic estimators, our findings further confirm that the Liljestrand and Zander method consistently outperforms other simple estimators, including those based on the Windkessel and Herd formulas. The Liljestrand and Zander estimator demonstrated the strongest associations with the thermodilution reference standard across multiple performance metrics, including the highest Spearman correlation and the lowest MAPE and MAE of the estimators examined. This aligns with prior reports and likely reflects the method’s superior ability to account for the nonlinear relationship between vascular compliance and arterial pressure [15]. Our results further reinforce the assertion that among straightforward physiologic approaches, the Liljestrand and Zander estimator is currently the preferred option for population-level CI estimation in the cardiac surgery setting.

Several important limitations should be acknowledged when interpreting these findings. First, a key limitation of the thermodilution-derived CI measurements serving as the CI reference standard for the study, was that details regarding the manufacturer and version of each device generating CI data was not available for analysis. While a multilevel model accounting for clustering by hospital may have partially addressed interhospital differences in thermodilution-based CI monitoring devices commonly used, specific device effects could not be fully disentangled. Similarly, granular data on clinician measurement technique or documentation practices were not available for study, introducing possible biases that may have been only partially mitigated by including the primary anesthesiologist as a random effect within the models developed. Second, our exclusive focus on adult cardiac surgical cases may restrict generalizability to other perioperative or critical care populations for which CI measurements may be clinically relevant. Third, the demographics of the cohort studied, consisting primarily of elderly White males, limits the generalizability of study findings to other underrepresented subgroups. Finally, whereas the study included multiple US hospitals, findings from our study may have limited generalizability to other cardiac surgical settings, such as smaller community hospitals within the US, or larger academic medical centers outside of the US. Nevertheless, the real-world, pragmatic nature of our multicenter data and the robust analytic approach provide a meaningful step forward toward broader, registry-based use of CI estimation models in cardiac surgery research.

## Conclusion

In conclusion, our study demonstrates that routinely collected physiologic data from intraoperative anesthesia records, when combined with patient and procedural factors, can be used to generate population-level estimates of cardiac index in adult cardiac surgical patients with modest error. However, the limitations to accurately classifying individual patients with normal versus low CI underscore the continued need for additional data (e.g. echocardiography) to guide individual clinical management. Nonetheless, these CI estimation models offer a tool for future observational studies investigating the role of CI-modifying interventions in influencing surgical outcomes potentially mediated by CI, with the potential to enhance registry-based research and quality improvement initiatives in cardiac surgery. Ongoing refinement and external validation will be essential to realize the full utility of such approaches.

## Supporting information

Online Resource 1

Online Resource 2

## Data Availability

The datasets involved in this study are defined as limited datasets per United States Federal Regulations and require execution of a data use agreement for transfer or use of the data. They are derived from data shared within the Multicenter Perioperative Outcomes Group (MPOG). The investigative team is able to share data securely and transparently conditional on: (i) receipt of a detailed written request identifying the requestor, purpose and proposed use of the shared data, (ii) use of a secure enclave for the sharing of personally identifiable information and (iii) the request is permissible within the confines of existing data use agreements executed between MPOG members.

## Acknowledgements

The authors gratefully acknowledge Robert Coleman, BS, (Department of Anesthesiology, University of Michigan Medical School, Ann Arbor, MI, USA) for his contributions in data acquisition and electronic search query programming for this project.

We thank the Multicenter Perioperative Outcomes Group (MPOG) Non-author Collaborators: Jie Cao, MPH (Joan and Irwin Jacobs Center for Health Innovation, University of California, San Diego, CA, USA); Allison Janda, MD (Department of Anesthesiology, University of Michigan Medical School, Ann Arbor, MI, USA); Donald Likosky, PhD (Department of Cardiac Surgery, University of Michigan Medical School, Ann Arbor, MI, USA); Robert B. Schonberger, MD, MHS, MHCDS (Department of Anesthesiology, Yale School of Medicine, New Haven, CT, USA); Robert Hawkins, MD, MSc (Department of Cardiac Surgery, University of Michigan Medical School, Ann Arbor, MI, USA); Michael Heung, MD, MS (Department of Internal Medicine - Nephrology Division, University of Michigan Medical School, Ann Arbor, MI, USA); Gorav Ailawadi, MD, MBA (Department of Cardiac Surgery, University of Michigan Medical School, Ann Arbor, MI, USA); Rahul Ladhania, PhD (Departments of Health Management and Policy, and Biostatistics, University of Michigan, Ann Arbor, MI, USA); Michael Sjoding, MD, MSc (Department of Internal Medicine, Pulmonary and Critical Care Medicine, University of Michigan Medical School, Ann Arbor, MI, USA); Sachin Kheterpal, MD, MBA (Department of Anesthesiology, University of Michigan Medical School, Ann Arbor, MI, USA);

The authors acknowledge the following funding sources for this project: The University of Michigan Medical School and the National Institute of Diabetes and Digestive and Kidney Diseases (NIDDK), National Heart, Blood, and Lung Institute (NHLBI), and the National Institute of General Medical Sciences (NIGMS) at the US National Institutes of Health.

## Notes

### Competing Interest Statement

Dr. Mathis reported receiving grants from the National Institute of Diabetes and
Digestive and Kidney Diseases (NIDDK) and the National Heart, Lung, and Blood Institute (NHLBI) of the National Institutes of Health (NIH) during the conduct of the study and Chiesi industry-sponsored research funding outside the submitted work. Dr Singh reported receiving grants from the NIDDK during the conduct of the study.

### Author Declarations

The IRBMED of the University of Michigan waived ethical approval for this work (HUM00206345).

