## Supplementary material for "Feasibility of Estimating Cardiac Indices Using Cardiac Surgery Anesthesia Records in a Multicenter Cohort": Online Resource 1: Online Resource 1. Supplemental Tables and Figures.docx

**Supplemental Table 1.** Number of cases and primary anesthesia attendings by study hospital. Participating health systems are Brigham and Women’s Hospital, Boston, Massachusetts; Massachusetts General Hospital, Boston, Massachusetts; Oregon Health and Science University, Portland, Oregon; University of Michigan Health System, Ann Arbor, Michigan; University of Washington Medical Center, Seattle, Washington; Washington University of St. Louis School of Medicine, St. Louis, Missouri; Yale New Haven Hospital, New Haven, Connecticut.

| Hospital | Pre-CPB Patients  n (%) | Post-CPB Patients  n (%) | Pre-CPB Primary Anesthesia Attending  n (%) | Pre-CPB Primary Anesthesia Attending  n (%) |
| --- | --- | --- | --- | --- |
| A | 1,219 (59) | 3,919 (70) | 35 (22) | 36 (18) |
| B | 598 (29) | 613 (11) | 45 (28) | 46 (23) |
| C | 73 (4) | 496 (9) | 29 (18) | 38 (19) |
| D | 105 (5) | 193 (3) | 22 (14) | 23 (11) |
| E | 22 (1) | 271 (5) | 10 (6) | 22 (11) |
| F | 33 (2) | 45 (1) | 15 (9) | 16 (8) |
| G | 4 (0) | 81 (1) | 3 (22) | 22 (11) |

### **Supplemental Table 2.** Correlation matrix for all cardiac index estimators. MAP = mean arterial pressure.

|  | MAP | Windkessel | Herd | Liljestrand and Zander |
| --- | --- | --- | --- | --- |
| MAP | 1.00 | 0.49 | 0.49 | 0.18 |
| Windkessel | 0.49 | 1.00 | 1.00 | 0.89 |
| Herd | 0.49 | 1.00 | 1.00 | 0.89 |
| Liljestrand and Zander | 0.18 | 0.89 | 0.89 | 1.00 |

### **Supplemental Table 3.** Confusion matrices for all cardiac index estimators in discriminating low CI (<2.2 L/min/m2) from normal or high CI (>=2.2 L/min/m2) compared to reference standard CI values from thermodilution. CI = cardiac index; CPB = cardiopulmonary bypass; MAP = mean arterial pressure.

|  | **Pre-CPB** | | | | | **Post-CPB** | | | |
| --- | --- | --- | --- | --- | --- | --- | --- | --- | --- |
| MAP |  |  | Predicted | |  |  |  | Predicted | |
|  |  |  | *Normal* | *Low* |  |  |  | *Normal* | *Low* |
|  | Actual | *Normal* | 608 | 317 |  | Actual | *Normal* | 3143 | 398 |
|  |  | *Low* | 484 | 645 |  |  | *Low* | 1583 | 494 |
| Windkessel |  |  | Predicted | |  |  |  | Predicted | |
|  |  |  | *Normal* | *Low* |  |  |  | *Normal* | *Low* |
|  | Actual | *Normal* | 605 | 320 |  | Actual | *Normal* | 3119 | 422 |
|  |  | *Low* | 445 | 684 |  |  | *Low* | 1486 | 591 |
| Herd |  |  | Predicted | |  |  |  | Predicted | |
|  |  |  | *Normal* | *Low* |  |  |  | *Normal* | *Low* |
|  | Actual | *Normal* | 605 | 320 |  | Actual | *Normal* | 3119 | 422 |
|  |  | *Low* | 445 | 684 |  |  | *Low* | 1486 | 591 |
| Liljestrand and Zander |  |  | Predicted | |  |  |  | Predicted | |
|  |  |  | *Normal* | *Low* |  |  |  | *Normal* | *Low* |
|  | Actual | *Normal* | 599 | 326 |  | Actual | *Normal* | 3094 | 447 |
|  |  | *Low* | 430 | 699 |  |  | *Low* | 1441 | 636 |

### **Supplemental Figure 1.** Bland-Altman plots for the Liljestrand and Zander estimator pre- and post-CPB. CI = cardiac index; CPB = cardiopulmonary bypass; MAP = mean arterial pressure.

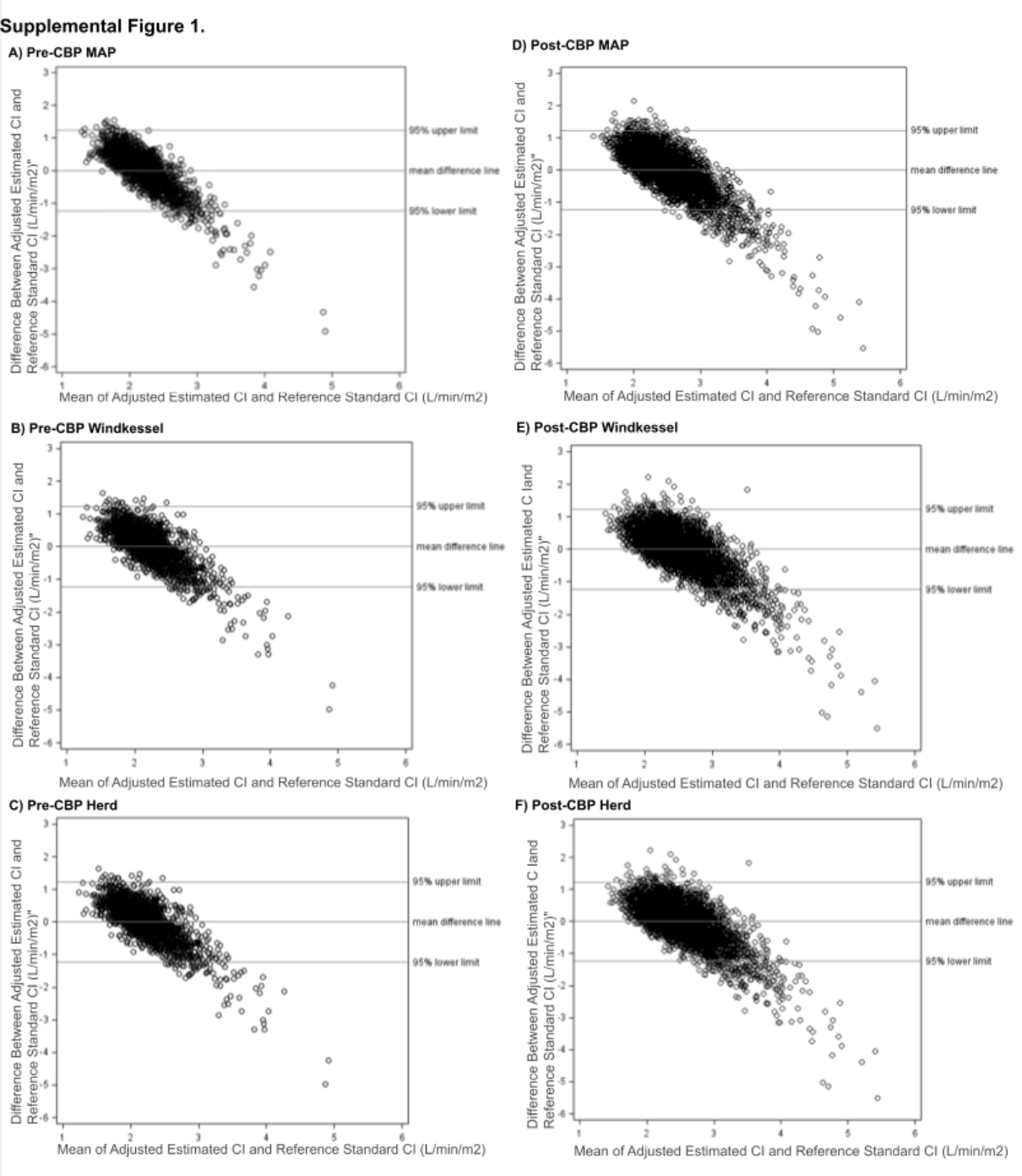
